# Clinical Utility of SPECT Neuroimaging in the Diagnosis and Treatment of Traumatic Brain Injury: A Systematic Review

**DOI:** 10.1101/2022.07.15.22277668

**Authors:** Michael Hanna, Jaclyn Herman, Bartosz Zawada, Christine Andraos, Oscar Karbi, Carina D’Souza, Atiemo Kessie, Getachew Mazengia, Sameer D’Souza

## Abstract

**Background:** The most common assessment modalities to determine the level of injury following a traumatic brain injury (TBI) includes computerized tomography (CT) scans and/or magnetic resonance imaging (MRI). Evidence is mixed as to whether single photon emission computed tomography (SPECT) is specific and accurate in identifying TBI.

**Objectives:** This study systematically assessed recent evidence of the clinical utility of SPECT in the diagnosis of TBI and examined the diagnostic accuracy of SPECT in TBI and its performance in comparison to other imaging modalities (e.g., CT and MRI).

**Methods:** PubMed, MEDLINE, and Embase databases were systematically searched for published articles from December 2012 to July 2022. Randomized controlled trials (RCTs) and observational studies published in English that used SPECT to evaluate patients with all severity of TBIs were eligible for inclusion. Titles and abstracts were screened, and 111 selected full-text articles were independently screened based on predefined inclusion/exclusion criteria (guided by the Preferred Reporting Items for Systematic Reviews and Meta-Analyses; PRISMA) and assessed for quality using the Newcastle-Ottawa Scale.

**Results:** Fourteen eligible studies, all observational, reporting location of lesions on brain SPECT were included, reporting data from 21632 participants of which 20,746 participants were from one study; the remaining 886 participants were from the remaining13 studies. The heterogeneity of the data precludes a meta-analysis. There was no consensus among experts from the thirteen smaller studies; however, the largest study indicated that the specificity of visual readings was 54%. In particular, abnormalities and brain perfusions may lead to false positives. Quantitative analysis theoretically increases the reliability of findings for brain SPECT, but error rates are unknown and not published.

**Conclusion:** There is a lack of evidence to support the clinical utility of brain SPECT for the diagnosis and treatment of TBI.

## Introduction

1. A traumatic brain injury (TBI) results from trauma to the brain, and is a major cause of disability and death (cite). Each, year it is estimated that approximately 69 million people suffer from a TBI worldwide, of which the majority of cases are mild (81%) or moderate (11%) [1]. Depending on the severity of the injury, those who sustain a TBI may face health problems that can range from short to long-term in duration. Because prompt proper management of a TBI can significantly alter the clinical course, especially within 48 h of the injury, neuroimaging techniques have become an important part of the diagnostic work up of such patients. Thus, diagnostic accuracy for TBI is critical for medical treatment planning in order to reduce common and potentially devastating clinical outcomes
2. While there are numerous definitions of TBI and there are ongoing debates within the scientific literature and among practicing professionals about the identification, signs, symptoms, diagnosis, and treatment of TBI of all severities, [2,3,6,7, 8, 9, 10] at this time, the most common assessment modality to determine the level of injury sustained after brain trauma includes two-dimensional modalities, computerized tomography (CT) scans and magnetic resonance imaging (MRI) (cite). Based on the results from the assessment, a treatment plan is carried out (e.g., medications, rest, rehabilitation, and/or surgery in severe cases). While MRI and CT imaging are the current gold standard to diagnose TBI (cite), other imaging techniques could be economically beneficial for healthcare systems worldwide and provide better outcomes for patients [18]. One such technique is single photon emission computed tomography (SPECT).

This advanced neuroimaging modality is a nuclear scan created by American researchers in 1963 that uses radioactive tracers to develop three-dimensional (D) images of blood flow [14]. The brain SPECT has undergone critical updates over the decades and newer machines often employs multi-headed cameras followed by quantitative analysis and attenuation to relay metabolic changes and physiological data [11,14]. The spatial resolution of brain SPECT images are lower compared to other similar scans such as positron emission tomography (PET) scan and functional magnetic resonance imaging (fMRI) [15,16]. Brain SPECT has been utilized to study neurological conditions and psychiatric disorders, which may include but not limited to: mood disorders, post-traumatic stress disorder (PTSD), obsessive compulsive disorder (OCD) [11,17].

There are contradictory findings in the scientific literature on brain SPECT and TBI. A brief overview on advanced neuroimaging techniques concluded that “there is insufficient evidence supporting the routine clinical use of advanced neuroimaging for diagnosis and/or prognostication at the individual patient level” [19]. Several systematic reviews on this topic have been published over the last several years [11,15,16,20,21], and most reviews, with the exception of Raji and colleagues (2014) [11], have been highly critical of the SPECT scan. A major criticism of the SPECT scan for TBI is that the results produced are substandard qualitative assessments between different regions of the brain, and attenuation of results are still less quantitatively accurate than other medical imaging modality such as PET scan. For example, Jacobs et al (1994) [23] and a follow-up study by the same group of researchers (1996) [24] reported poor results from the SPECT scan for TBI after a 3-month follow-up period, which is when some signs and symptoms of minor head injury have already diminished. However, after a 12-month follow-up assessment, the sensitivity (100%), specificity (85%), negative predictive value (100%), and positive predictive value (83%) of brain SPECT for TBI were remarkably high. As such, it is questionable whether the findings from the SPECT scan are post-injury evidence or if these are incidental findings that are indicative of confounding factors such as separate medical conditions.

The most comprehensive systematic review of medical imaging technology to date was conducted by Amyot and colleagues (2015) [15], and noted that although the results are promising to support the clinical utility of SPECT scan for?. In contrast, a systematic review by Raji et al. (2014) [11] regarding the clinical utility of brain SPECT in the early? diagnosis and treatment of TBI cited high levels of sensitivity and specificity. Although the authors reported level IIA evidence, it should be noted that few studies utilized control groups or rigorous scientific methodology such as double-blind randomization. Furthermore, it is a logical fallacy to suggest *post hoc ergo propter hoc* (“after this, therefore because of this”), and thus, the clinical history and evidence-based decision-making are necessary to investigate whether post-injury findings are directly related to an event or accident. In order for the wider scientific community and healthcare industry to endorse the use of brain SPECT for TBI, the generalizability of the results of this medical imaging modality for different cohorts must be confirmed. Therefore, an updated review of this study is necessary to determine if the findings from the previous review are replicable and provide additional insights by including the latest empirical evidence available on this topic.

The purpose of this study was to update the systematic review by Raji and colleagues (2014) [11] and to evaluate the clinical utility of SPECT in the diagnosis and treatment of TBI, ranging from mild to severe The PICO statement and methods remained consistent from the prior study so current findings could extrapolate from previous results. Thus, this updated review aims to answer if abnormalities on brain SPECT could be utilize to diagnose and treat individuals with TBI in clinical settings in comparison to other brain imaging modalities. A secondary analysis was also conducted to identify possible associations between SPECT abnormalities and neuropsychological and neurological outcomes.

## Methods

This review followed protocols outlined in the PRISMA 2020 guidelines [25] and was submitted to PROSPERO for approval (ID: CRD42021276772). This updated review closely followed the methods provided in the previous systematic review conducted by Raji and colleagues (2014) [11]. However, the current authors decided to broaden the search strategy to comprehensively explore all possible resources and find all records related to this topic.

### Search Strategy

PubMed, MEDLINE (Ovid), Embase (Ovid), Google Scholar, and citation searching were utilized to identify relevant articles published in English between December 2012 to July 2021, in accordance with the previous date range of 1983 to November 2012 from the prior systematic review. The MESH terms used to search the databases are presented in Chart 1.

**Chart 1.**
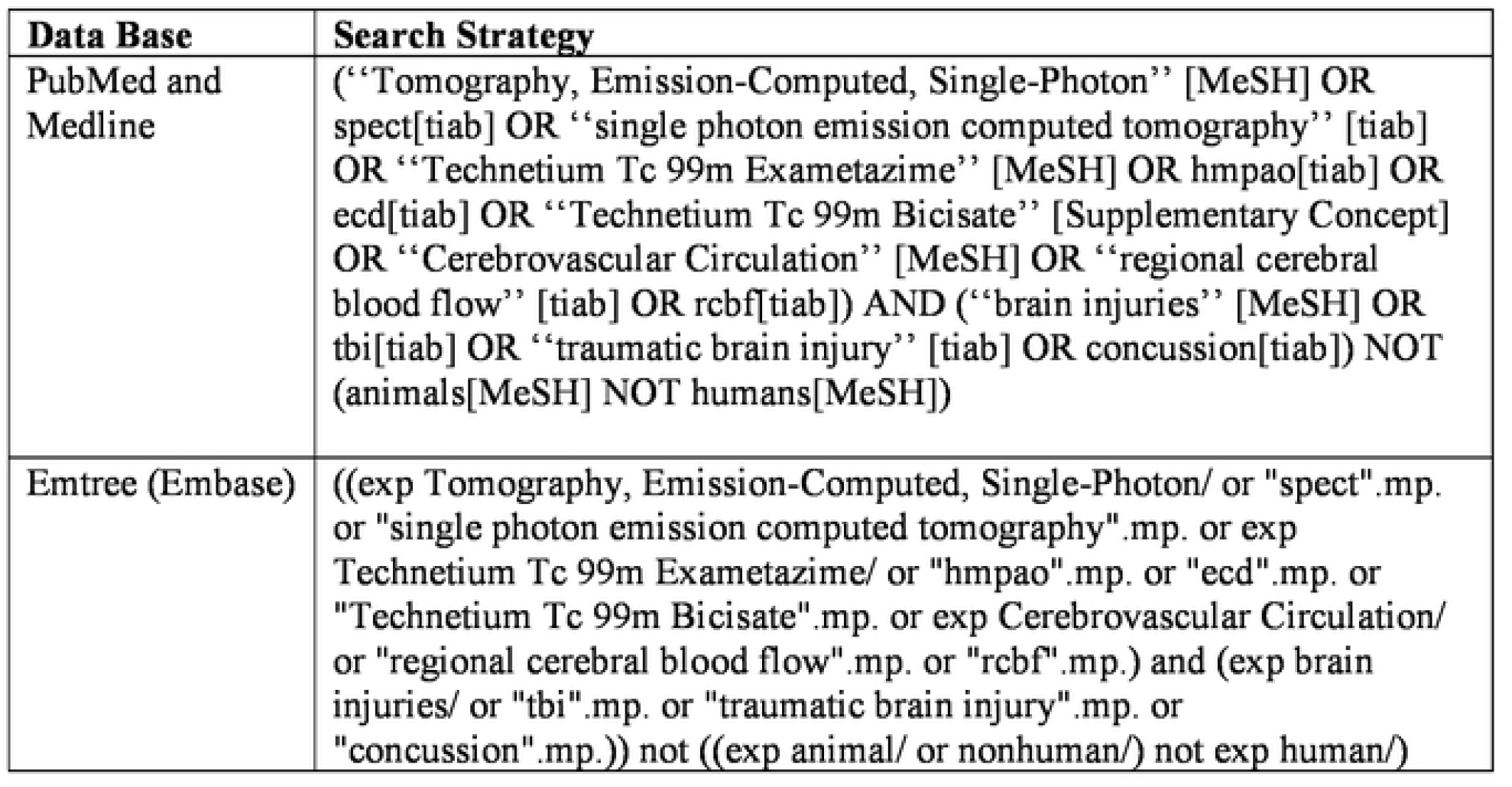

### Study Selection

The inclusion criteria were: (a) Randomized controlled trials (RCTs) and observational studies, including longitudinal, cross-sectional, and cohort studies that used brain SPECT to evaluate patients with TBI; (b) full-text peer reviewed studies published with a description of SPECT abnormalities; and (c) articles in English. The exclusion criteria were: (a) case studies and articles outside the specify date range; (b) studies without description of SPECT abnormalities; and (c) articles in foreign languages. Three reviewers participated in the identification of records, title and abstract screening, reviewing of full-text peer reviewed articles, and data extraction and analysis process.

### Data Extraction and Quality Assessment

Search results from electronic databases were uploaded in Mendeley and charted in Excel. Duplicates were automatically removed and examined for accuracy by one reviewer. Titles and abstracts were independently screened by two reviewers against inclusion and exclusion criteria. A third reviewer resolved any disagreement between other investigators. Only relevant articles were included for full-text assessment and data extraction. Abnormalities on brain scans, results from neuropsychological/neurological evaluations, and severity of TBI were extracted for analysis. Additionally, study setting and demographic factors were noted. One reviewer assessed the quality of all eligible studies utilizing a standard quality assessment tool, the Newcastle-Ottawa Scale [26,27]. A second reviewer checked the results and consultation with a third reviewer occurred if there is any disagreement. A narrative synthesis described whether brain SPECT is specific in identifying TBI. Descriptive statistics summarized the findings and post-test probabilities. Measures of diagnostic accuracy were extracted for analysis if the measures were included in the report (sensitivity, specificity, and negative/positive predictive values). Statistical analysis was calculated using the appropriate packages on the latest version of R via R Studio.

### Amendment to Protocol

Due to a lack of responsiveness for edits, one author was removed

## Results

Title and abstract screening included 2235 articles found on electronic databases (Fig 1): PubMed (*n* = 738), Ovid Medline (*n* = 671), Ovid Embase (*n* = 843), and other sources (*n* = 1). Two hundred and eleven duplicates were removed and an additional 1915 articles were excluded following title and abstract screening. A total of 111 full-text peer reviewed articles were analyzed, of which 14 observational studies were selected for further analysis [28–41]. The inter-rater reliability for the article selection process was calculated to be over 80%, which is considered acceptable levels (*κ* = 0.87, *p* < .05). It should be noted that no RCT was found and the heterogeneity of the data precludes conducting a meta-analysis. The combined sample size for this study is 21,416 persons with TBI of all severity.

**Fig 1.**
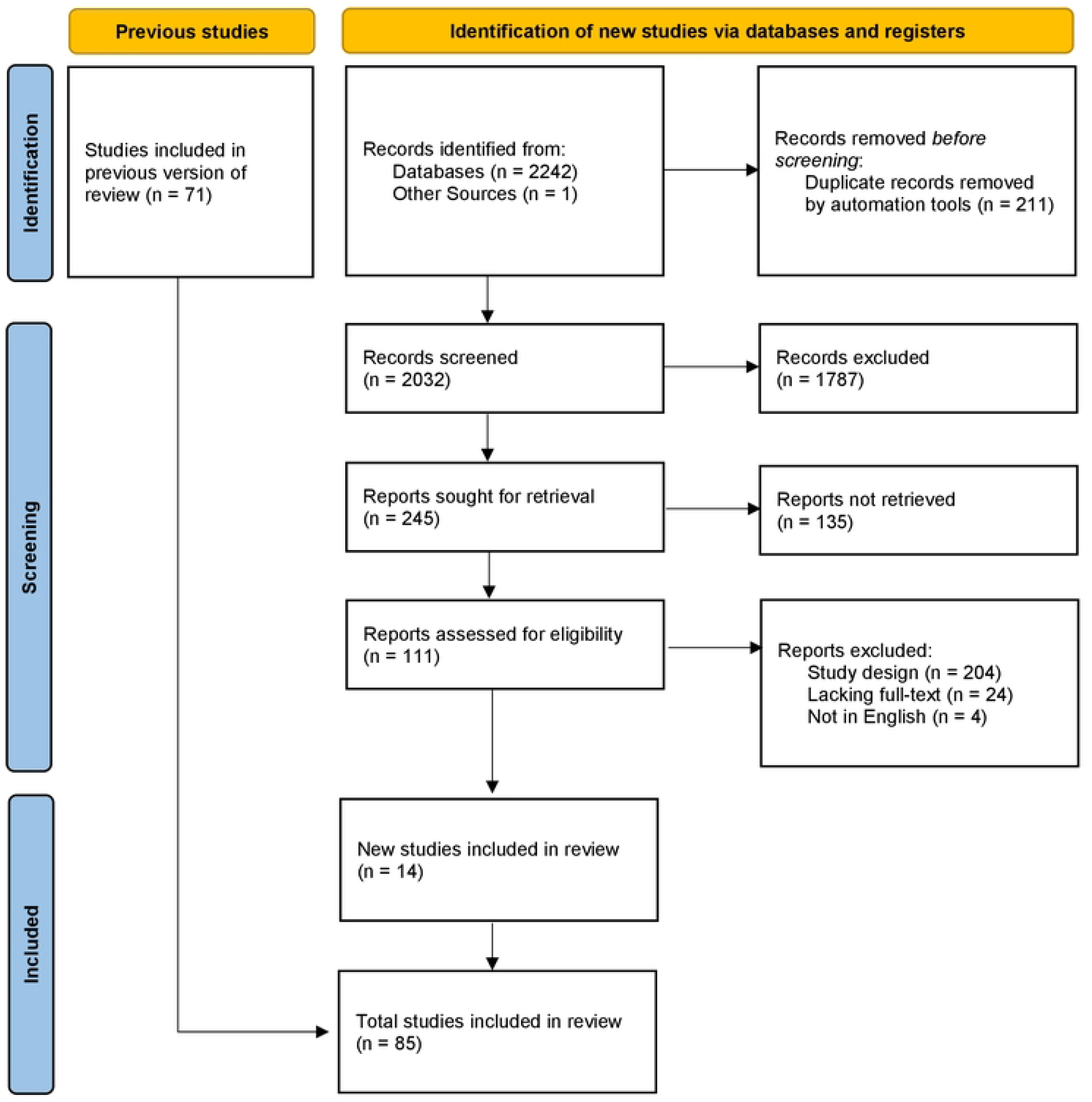
Flowchart of Article Selection for Updated Systematic Review. *Note* n = number of records; Adapted from the PRISMA 2020 Statement An Updated Guideline for Reporting Systematic Reviews [25].

**Fig 2.**
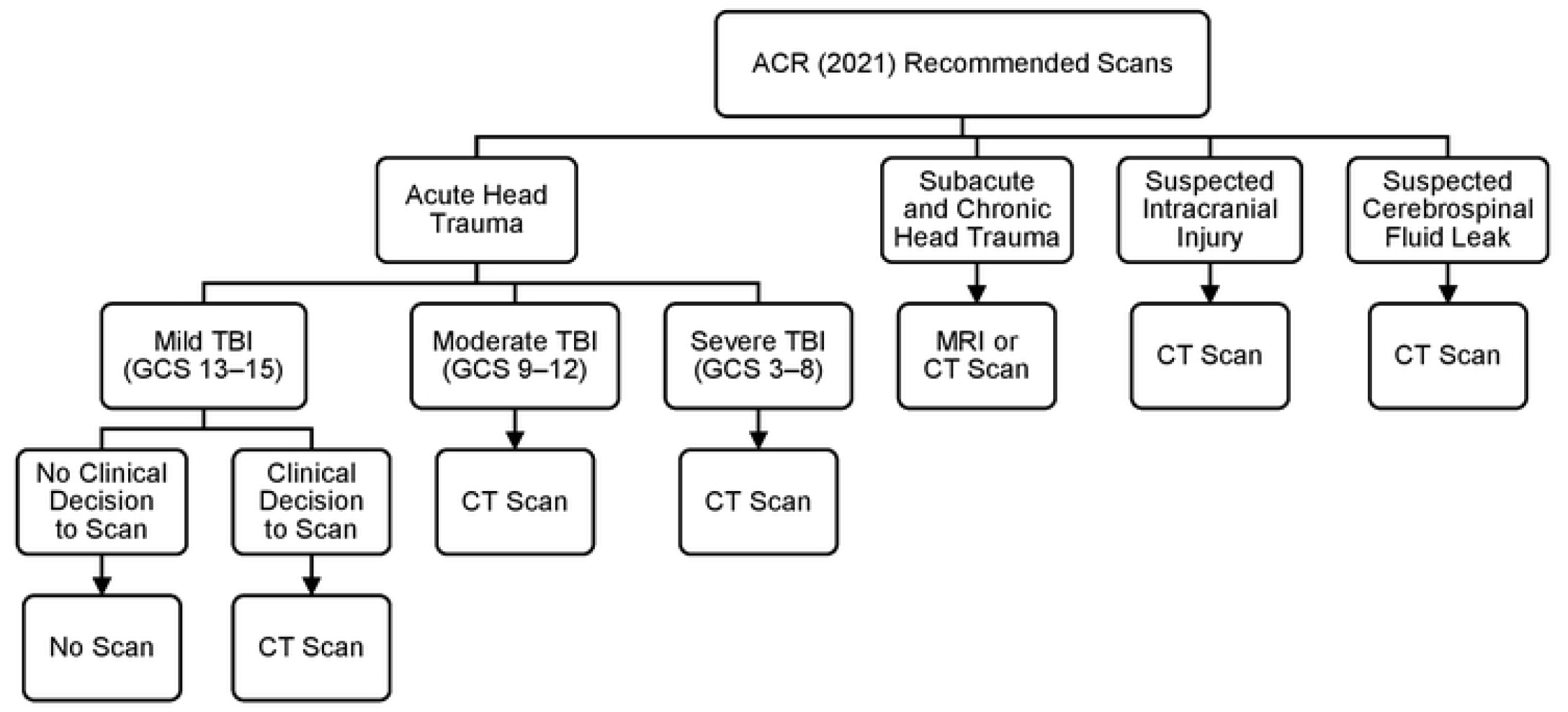
American College of Radiology Recommended Scans for Head Trauma. *Note:* TBI = traumatic brain injury; GCS = Glasgow Coma Scale; CT= computed tomography; MRI = magnetic resonance imaging; Adapted from the American College of Radiology (ACR) Appropriateness Criteria Head Trauma: 2021 Update [13].

### Longitudinal Cohort Studies

Three longitudinal cohort studies investigated SPECT scan for TBI, which included 405 participants in total (146/405 participants were healthy controls), and one study did not include a control group for comparison (Table 1) [28]. All three studies were conducted in different locations (United States, Canada, and Denmark) and different tracers were used to inject participants for each study (HMPAO, ^99^mTc-ECD, and ^123^I-CLINDE). The majority reported using a scanner with a triple-headed camera system (66%) and only one study reported using CT scan (33%) and MRI for comparison (33%). Concussion or mTBI were the focus of each study (100%) and the majority reported lesion locations associated with the frontal lobe (100%), parietal lobe (66%), temporal lobe (66%), cerebellum (66%), and occipital lobe (33%). One of three studies compared professional athletes to healthy controls and found that all regions of the brain had indications of lower cerebral perfusion, which also correlated with psychological testing [29]. One study reported that there was no significant association between visual inspection of brain SPECT scan and clinical outcomes [28]. Similarly, researchers also found that no significant difference was detected after statistical corrections were performed [30]. Neuropsychological outcomes associated with post-concussion syndrome were investigated in two studies and both studies found no correlation [28,30].

**Table 1.** Summary of Longitudinal Cohort Studies.

| Study (Year) | N, Start /Follow Up | Age | Male Gender (%) | Lesion Localization | Follow Up, months | NOS Score |
| --- | --- | --- | --- | --- | --- | --- |
| Amen (2016) [29] | 285 | 52 | 100 | F/T/P/O/C | Not mentioned | 7 |
| Ebert (2019) [30] | 36/35 | 42 | 56 | F/T/C | 3–4 | 7 |
| Romero (2015) [28] | 84/49 | 32 | 65 | F/P | 12 | 6 |

*Note:* N = sample size; F = frontal lobe; T = temporal lobe; P = parietal lobe; O = occipital lobe; C = cerebellum; NOS = Newcastle-Ottawa Scale.

### Observational Studies

There were 14 observational studies that examined brain SPECT and TBI (Table 2). All studies reported positive findings between SPECT scan and TBI. However, there were mixed findings for some conditions such as post-concussion syndrome and mood disorders [31,32]. In total, there were 21632 participants included in these studies but 20,746 participants were included in a single study conducted by Amen et al. (2015) [33]; and furthermore, Raji et al. (2015) [34] extrapolated data using the same sample of participants. With the exception of the two aforementioned studies, there were only 265 participants in total from the additional 12 prospective cohort studies that were found and the majority had an insufficient number of participants to achieve optimal statistical power due to the small sample size (7 – 73 participants in each study) [31,32,35–41]. Four studies investigated all severities of TBI (36%), five studies focused on mTBI (45%), one study recruited those with severe TBI (9%), and one study did not define the severity of TBI (9%). Most studies only utilized SPECT scan (i.e., no comparison imaging, 55%), five studies used MRI as the comparison imaging (45%), and lastly, PET scan (9%) and CT scan (9%) were each included in only one study that also used MRI as a comparison modality. Different nuclear tracers were used for each study and method of analysis did not conform to a single protocol. Time of SPECT was normally months after initial injury (55%) and lesion detection were reportedly not seen on comparison imaging (100%). Injury was primarily found on the frontal lobe (100%), temporal lobe (64%), parietal lobe (55%), cerebellum (55%), and least of all, the occipital lobe (36%). Several studies found that SPECT scan can clinically differentiate TBI from PTSD between the general population, veterans, and healthy controls [33,34]. The only study that compared PET and SPECT scan determined that PET scan is better at detecting lesions than brain SPECT for TBI [35]. However, brain abnormalities were often identified on brain SPECT independent of results indicated from structural scans such as CT scan and MRI [32,35–37].

**Table 2.** Summary of Observational Studies.

| Study (Year) | TBI Type | Sample Size | Comparison Imaging | Time of SPECT | Lesion Localization | Lesion Detection Not Seen on Comparison Imaging |
| --- | --- | --- | --- | --- | --- | --- |
| Abiko (2017) [35] | All severities | 7 | PET and MRI | Months to Years | F/T/P/C | Yes |
| Amen (2015) [33] | All severities | 20746 | SPECT Only | Not mentioned | F/T/P/O/C | Not applicable |
| Crider (2018) [36] | Mild | 12 | CT and MRI | Months after TBI | F/T | Yes |
| Khalili (2020) [39] | Not defined | 12 | SPECT Only | Months to Years | F/P | Not applicable |
| Nakagawara (2013) [40] | Mild | 17 | MRI | Months to Years | F | Yes |
| Newberg (2014) [38] | Mild | 45 | SPECT Only | Months after TBI | F/T/P/O/C | Not applicable |
| Raji (2015) [34] | All severities | 196 | SPECT Only | Not mentioned | F/C | Not applicable |
| Romero (2014) [31] | Mild | 67 | SPECT Only | Weeks after TBI | F/O | Not applicable |
| Uruma (2013) [37] | Mild | 73 | MRI | Months to Years | F/T/P/C | Yes |
| Winter (2015) [32] | All severities | 14 | MRI | Days after TBI | F/T/P/O/C | Yes |
| Womack (2020) [41] | Moderate, Severe | 18 | SPECT Only | Weeks after TBI | F/T | Not applicable |

*Note:* TBI = traumatic brain injury; SPECT = single photon emission computed tomography;
PET = Positron emission tomography; MRI = magnetic resonance imaging; F = frontal lobe; T =
temporal lobe; P = parietal lobe; O = occipital lobe; C = cerebellum.

### Diagnostic Accuracy

Previous results from the systematic review conducted by Raji and colleagues (2104) [11] highlighted exemplary findings from three longitudinal cohort studies that reported measures of diagnostic accuracy (sensitivity, specificity, and predictive values). In particular, the authors emphasized the importance of follow-up evaluations to identify and diagnose TBI on SPECT scans. As mentioned, the results from Jacobs and colleagues (1994) [23] and a follow-up assessment (1996) [24] described remarkably high sensitivity (100%) and specificity (85%) for brain SPECT and TBI. In addition, the sensitivity and specificity of SPECT were reported in four recent cohort studies. There are indications that poor neurological findings (e.g., cognitive dysfunctions and memory/attention deficits) were correlated with brain SPECT for mTBI (sensitivity ≥ 73% and specificity ≥ 75%) [37]. In a cross-sectional study, Raji and colleagues (2015) [34] described that brain SPECT can distinguish PTSD from all severities of TBI, but the results are only moderately reflected in the calculated sensitivity (≥ 80%) and specificity (≥ 64%) values. In comparison to previously reported results, two studies reported lower sensitivity (≥ 67%) and most found similar specificity levels (≥ 54%) as illustrated in Table 3. The largest cohort study (*N* = 20,746 participants) involving SPECT for TBI reported specificity was 54% at rest and on-task for visual readings of scans [33]. This coincides with findings from a longitudinal study by Romero and colleagues (2015) [28] who described there were no association between visual inspections in radiological reports with clinical outcomes for TBI. Although quantitative analysis was included in most recent studies, none reported error rates and methods of analysis varied between studies. Furthermore, predictive values were not reported in the results of recent studies found for this review article, and only a single study reported predictive models for sensitivity and specificity measures, which were 86% and 80% respectively among professional athletes who experienced sport-related concussions [29].

**Table 3.** Sensitivity and Specificity of Brain SPECT in Peer Reviewed Publications.

| Study (Year) | Sensitivity | Specificity |
| --- | --- | --- |
| Jacobs (1996) [24] | 78% – 100% | 53% – 85% |
| Uruma (2013) [37] | 73% – 100% | 75% – 83% |
| Raji<br>(2015) [34] | 80% – 92% | 64% – 85% |
| Amen<br>(2015) [33] | 67% – 100% | 54% – 100% |
| Amen<br>(2016) [29] | 86% – 90% | 80% – 87% |

*Note:* The sensitivity and specificity data were reported in the article. The data from Jacobs et al.
(1996) [24] was included in the systematic review by Raji et al. (2014) [11].

## Discussion

The overall results of this study indicated that brain SPECT could still be considered an experimental procedure that lacks scientifically sound evidence. Longitudinal findings associated with sport-related concussions among professional athletes [29] and cross-sectional results examining those with PTSD and TBI [34] could be promising but the results may not be generalizable for different patient population. Additional studies with larger samples are required to confirm the findings from the current results. Two studies with large samples utilized identical retrospective patient evaluations; and consequently, the study designs lack proper study protocols such as randomization and blinding [33,34]. As such, limited inferences can be made from the results of these studies. Most notably, PET scan outperformed SPECT scan at detecting brain lesions [35], and several studies found that brain SPECT did poorly to differentiate mood disorders and post-concussion syndrome [28,30–32]. Furthermore, there is a lack of evidence to indicate the usefulness of SPECT scan in the diagnosis and treatment of moderate traumatic brain injury (studies rarely investigate this classification of brain injury). Although more recent literature provided better evidence to support contemporary quantitative analysis techniques for SPECT scan, certain factors such as error rates of this modality are still unknown and the lack of quality in the resulting images is undisputable [11,14–16,42]. In particular, abnormalities and brain perfusions could be detected based on visual inspection, but may lead to false positives without mathematical models and corrections. Quantitative analysis using normal database theoretically increases the reliability of findings for brain SPECT, but as mentioned the error rates are still unknown and not published, and therefore results derived from SPECT scans are not infallible. Raji and Henderson (2018) [42] published a brief narrative update to the systematic review on clinical utility of SPECT in the diagnosis and treatment of traumatic brain injury. The authors compared SPECT, PET scans, and functional MRI (fMRI) and provided an overview of issues relating to the SPECT scan. Key points of this article are that the SPECT scan is still clinically useful for diagnosis of TBI because comparison modality such as PET scan lacks large studies and fMRI requires better post-processing procedures. However, the authors also mentioned that SPECT, PET, and fMRI are all “advanced neuroimaging techniques” that are still “areas of active research but are not considered routine clinical practice at this time.” (in accordance with the ACR 2016 guideline for head trauma) [12]. Therefore, the general consensus is that these medical imaging modalities are still within the realm of experimental research and emphasis should be placed on clinical decisions when diagnosing patients with complex issues such as TBI.

Further complicating this topic is the lack of a widely acceptable clinical practice guideline. Although it has been argued that the lower cost of SPECT scan could help provide “maximal benefit to patient care,” [42] the potential legal implications are unclear when brain SPECT findings are presented during litigation. It is well-documented in the literature that TBI cases are multifaceted in nature and involved stakeholders from different professional backgrounds [43,44]. A universally accepted clinical practice guideline will provide evidence of evaluation and treatment to clarify issues presented during civil litigation [43]. However, an unpublished guideline may further create confusion within the medicolegal contexts. The latest guideline for brain perfusion SPECT by the Canadian Association of Nuclear Medicine (CANM) included similar claims about the prognostic value of brain SPECT that was endorsed by the EANM over a decade earlier [17], and as such adherence to the recommendations is warranted with abundance of caution. Additionally, SPECT scan would not meet the Daubert Standard for expert’s scientific testimony because its “potential error rate” is unknown (Criterion 3) and it has not “attracted widespread acceptance within relevant scientific community” (Criterion 5) [44]. In comparison to the widely accepted use of CT scan and MRI to diagnose TBI, brain SPECT is not supported by current health regulations or experts in the field. Medical diagnostic imaging must be precise and accurate because these are common tools physicians routinely rely on during the course of their daily practice.

The bottom line is that clinicians should interpret the SPECT findings in the context of clinical history. This view is consistent with correspondence between Adinoff and Devous (2010) [46] and Amen (2010) [47], who agreed that the evidence base to support the application of SPECT for diagnosis and treatment without the consideration of other sources of data is insufficient. As a rule of thumb in clinical medicine, imaging, laboratory, and radiological findings need clinical correlation. As noted by Amen (2010) [47], “Thoughtful clinicians would never use SPECT in isolation. That is not how imaging is or should be practiced.”

### Limitations and Future Directions

Adherence to the protocols from the previous systematic review was critical and this could have potentially limited the scope of this article (e.g., further investigation regarding different mental health issues beyond those associated with TBI) [48]. In addition, lower level of evidence such as case reports were not considered because level IIA evidence was required to achieve the same standards set by the previous authors. The current findings only included a narrative synthesis due to the varying methods of data collection (e.g., different tracers were injected into patients such as 99mTc-ECD or 99mTc-HMPAO) from each study. Additionally, it is unclear if the study setting were consistent (i.e., more details are needed for further analysis), which could have also potentially impacted the results of the imaging when performed under different circumstances. It is salient to note that the participants cognitive state (fatigue/alert) and demographic factors such as age (youth/adult/elderly) during time of examination could have impacted the results of the metabolic scan. For instance, cerebral microbleeds or microhemorrhages are increasingly recognized as natural occurrences due to aging, but they have also been detected on medical images for TBI, which could potentially cause false-positives [49,50]. However, the small sample size from most studies found in this review would have made it highly difficult to further analyze possible confounding factors that could have impacted the findings of the SPECT scan. Although this systematic review did not include a meta-analysis, more research on this topic is required before additional statistical analysis can be performed. Future studies must utilize larger sample size and scientifically rigorous methodology (i.e., double-blind RCTs) to reduce risk of bias and enhance the strength of evidence available to support clinical utility of SPECT for diagnosis of TBI. Lastly, a detailed report regarding brain SPECT error rates is required before further inferences about its clinical utility are determined.

## Conclusion

Recent findings on this topic helps provide clarity on whether SPECT is specific in identifying TBI even without typical structural imaging such as CT and MRI. In conclusion, there is no consensus among experts about the clinical utility of brain SPECT. This “advance” medical imaging modality is currently not the gold standard diagnostic scan to investigate TBI. CT Scan and MRI are widely used and acceptable diagnostic imaging for TBI. PET scan is another advance neuroimaging modality that consideration should be given because it can outperform brain SPECT to detect abnormalities and lesions. Lastly, additional research is required to understand the intricacies regarding abnormalities on brain SPECT and conditions with clinical evidence similar to TBI. At the present, there is a lack of evidence to support the clinical utility of brain SPECT for the diagnosis and treatment of TBI in daily practice and healthcare settings. Therefore, diagnosis of TBI should be supported by evidence-based clinical decisions.

## Data Availability

N/A

## References

1. Dewan MC, Rattani A, Gupta S, Baticulon RE, Hung YC, Punchak M, et al. Estimating the global incidence of traumatic brain injury. J Neurosurg. 2019;130(4):1080–97. doi:10.3171/2017.10.JNS17352

2. Schneider KJ, Leddy JJ, Guskiewicz KM, Seifert T, McCrea M, Silverberg ND, et al. Rest and treatment/rehabilitation following sport-related concussion: A systematic review. Br J Sports Med. 2017;51(12):930–4. doi:10.1136/bjsports-2016-097475

3. Silverberg ND, Iverson GL, Arciniegas DB, Bayley MT, Bazarian JJ, Bell KR, et al. Expert panel survey to update the American Congress of Rehabilitation Medicine definition of mild traumatic brain injury. Arch Phys Med Rehabil. 2021;102(1):76–86. doi:10.1016/j.apmr.2020.08.022

4. Amen DG, Newberg A, Thatcher R, Jin Y, Wu J, Keator D, et al. Impact of playing American professional football on long-term brain function. J Neuropsychiatry Clin Neurosci. 2011;23(1):98–106. doi:10.1176/appi.neuropsych.23.1.98

5. DOD (Department of Defense). Psychological Health and Traumatic Brain Injury Research Program (PH/TBIRP); 2010. Available from: https://cdmrp.army.mil/phtbi/pbks/phtbipbk2011.pdf

6. Godoy DA, Rubiano A, Rabinstein AA, Bullock R, Sahuquillo J. Moderate traumatic brain injury: The grey zone of neurotrauma. Neurocrit Care. 2016;25(2):306–19. doi:10.1007/s12028-016-0253-y

7. Tator CH. Concussions and their consequences: Current diagnosis, management and prevention. CMAJ. 2013;185(11): 975–979. doi:10.1503/cmaj.120039

8. CDC (Centers for Disease Control and Prevention). Traumatic brain injury and concussion; 2021. Available from: https://www.cdc.gov/traumaticbraininjury/index.html

9. APA (American Psychiatric Association). Diagnostic and Statistical Manual of Mental Disorders, Fifth Edition (DSM-5). Arlington, VA: American Psychiatric Association; 2013.

10. Kay T, Harrington DE, Adams R, Anderson T, Berrol S, Cicerone K, et al. Definition of mild traumatic brain injury. J Head Trauma Rehabil. 1993;8(3):86–7.

11. Raji CA, Tarzwell R, Pavel D, Schneider H, Uszler M, Thornton J, et al. Clinical utility of SPECT neuroimaging in the diagnosis and treatment of traumatic brain injury: A systematic review. PLoS One. 2014;9(3):1–10. doi:10.1371/journal.pone.0091088

12. Shetty VS, Reis MN, Aulino JM, Berger KL, Broder J, Choudhri AF, et al. ACR appropriateness criteria head trauma. J Am Coll Radiol. 2016;13(6):668–79. doi:10.1016/j.jacr.2016.02.023

13. Shih RY, Burns J, Ajam AA, Broder JS, Chakraborty S, Kendi AT, et al. ACR appropriateness criteria head trauma: 2021 update. J Am Coll Radiol. 2021;18(5):S13–36. doi:10.1016/j.jacr.2021.01.006

14. Hutton BF. The origins of SPECT and SPECT/CT. Eur J Nucl Med Mol Imaging. 2014;41(S3–16). doi:10.1007/s00259-013-2606-5

15. Amyot F, Arciniegas DB, Brazaitis MP, Curley KC, Diaz-Arrastia R, Gandjbakhche A, et al. A review of the effectiveness of neuroimaging modalities for the detection of traumatic brain injury. J Neurotrauma. 2015;32(22):1693–721. doi:10.1089/neu.2013.3306

16. Lee AL. Advanced imaging of traumatic brain injury. Korean J Neurotrauma. 2020;16(1):3–17. doi:10.13004/KJNT.2020.16.E12

17. Kapucu ÖL, Nobili F, Varrone A, Booij J, Vander Borght T, Någren K, et al. EANM procedure guideline for brain perfusion SPECT using 99mTc-labelled radiopharmaceuticals. Eur J Nucl Med Mol Imaging. 2009;36(12):2093–102. doi:10.1007/s00259-009-1266-y

18. Hricak H, Abdel-Wahab M, Atun R, Lette MM, Paez D, Brink JA, et al. Medical imaging and nuclear medicine: A Lancet Oncology Commission. Lancet Oncol. 2021;22(4):136–72. doi:10.1016/S1470-2045(20)30751-8

19. Wintermark M, Sanelli PC, Anzai Y, Tsiouris AJ, Whitlow CT, Druzgal TJ, et al. Imaging evidence and recommendations for traumatic brain injury: Advanced neuro- and neurovascular imaging techniques. Am J Neuroradiol. 2015;36(2):1–11. doi:10.3174/ajnr.A4181.

20. Belanger HG, Vanderploeg RD, Curtiss G, Warden DL. Recent neuroimaging techniques in mild traumatic brain injury. J Neuropsychiatry Clin Neurosci. 2007;19(1):5–20. doi:10.1176/appi.neuropsych.19.1.5

21. Davalos DB, Bennett TL. A review of the use of single-photon emission computerized tomography as a diagnostic tool in mild traumatic brain injury. Appl Neuropsychol. 2002;9(2):92–105. doi:10.1207/S15324826AN0902_4

22. Stiell IG, Clement CM, Rowe BH, Schull MJ, Brison R, Cass D, et al. Comparison of the Canadian CT Head Rule and the New Orleans Criteria in patients with minor head injury. J Am Med Assoc. 2005;294(12):1511–8. doi: 10.1001/jama.294.12.1511

23. Jacobs A, Put E, Ingels M, Bossuyt A. Prospective evaluation of technetium-99m-HMPAO SPECT in mild and moderate traumatic brain injury. J Nucl Med. 1994;35(6):942–7.

24. Jacobs A, Put E, Ingels M, Put T, Bossuyt A. One-year follow-up of technetium-99m-HMPAO SPECT in mild head injury. J Nucl Med. 1996;37(10):1605–9.

25. Page MJ, McKenzie JE, Bossuyt PM, Boutron I, Hoffmann TC, Mulrow CD, et al. The PRISMA 2020 statement: An updated guideline for reporting systematic reviews. BMJ. 2021;372. doi:10.1136/bmj.n71

26. Farrah K, Young K, Tunis MC, Zhao L. Risk of bias tools in systematic reviews of health interventions: An analysis of PROSPERO-registered protocols. Syst Rev. 2019;8(1):1–9. doi:10.1186/s13643-019-1172-8

27. Wells GA, Shea B, O’Connell D, Peterson J, Welch V, Losos M, et al. The Newcastle-Ottawa Scale (NOS) for assessing the quality of nonrandomised studies in meta-analyses. Ottawa: Ottawa Hospital Research Institute; 2018. Available from: http://www.ohri.ca/programs/clinical_epidemiology/oxford.asp

28. Romero K, Lobaugh NJ, Black SE, Ehrlich L, Feinstein A. Old wine in new bottles: Validating the clinical utility of SPECT in predicting cognitive performance in mild traumatic brain injury. Psychiatry Res Neuroimaging. 2015;231(1):15–24. doi:10.1016/j.pscychresns.2014.11.003

29. Amen DG, Willeumier K, Omalu B, Newberg A, Raghavendra C, Raji CA. Perfusion neuroimaging abnormalities alone distinguish national football league players from a healthy population. J Alzheimers Dis. 2016;53(1):237–41. doi:10.3233/JAD-160207

30. Ebert SE, Jensen P, Ozenne B, Armand S, Svarer C, Stenbaek DS, et al. Molecular imaging of neuroinflammation in patients after mild traumatic brain injury: A longitudinal (123) I-CLINDE single photon emission computed tomography study. Eur J Neurol. 2019;26(12):1426–32. doi:10.1111/ene.13971

31. Romero K, Black SE, Feinstein A. Differences in cerebral perfusion deficits in mild traumatic brain injury and depression using single-photon emission computed tomography. Front Neurol. 2014;5:1–8. doi:10.3389/fneur.2014.00158

32. Winter C, Bell C, Whyte T, Cardinal J, Macfarlane D, Rose S. Blood-brain barrier dysfunction following traumatic brain injury: Correlation of K(trans) (DCE-MRI) and SUVR (99mTc-DTPA SPECT) but not serum S100B. Neurol Res. 2015;37(7):599–606. doi:10.1179/1743132815Y.0000000018

33. Amen DG, Raji CA, Willeumier K, Taylor D, Tarzwell R, Newberg A, et al. Functional neuroimaging distinguishes posttraumatic stress disorder from traumatic brain injury in focused and large community datasets. PLoS One. 2015;10(7):1–22. doi:10.1371/journal.pone.0129659

34. Raji CA, Willeumier K, Taylor D, Tarzwell R, Newberg A, Henderson TA, et al. Functional neuroimaging with default mode network regions distinguishes PTSD from TBI in a military veteran population. Brain Imaging Behav. 2015;9(3):527–34. doi:10.1007/s11682-015-9385-5

35. Abiko K, Ikoma K, Shiga T, Katoh C, Hirata K, Kuge Y, et al. I-123 iomazenil single photon emission computed tomography for detecting loss of neuronal integrity in patients with traumatic brain injury. EJNMMI Res. 2017;7(1):1–10. doi:10.1186/s13550-017-0276-1

36. Crider T, Eng D, Sarkar PR, Cordero J, Krusz JC, Sarkar SN. Microvascular and large vein abnormalities in young patients after mild head trauma and associated fatigue: A brain SPECT evaluation and posture dependence modeling. Clin Neurol Neurosurg. 2018;170:159–64. doi:10.1016/j.clineuro.2018.05.019

37. Uruma G, Hashimoto K, Abo M. A new method for evaluation of mild traumatic brain injury with neuropsychological impairment using statistical imaging analysis for Tc-ECD SPECT. Ann Nucl Med. 2013;27(3):187–202. doi:10.1007/s12149-012-0674-4

38. Newberg AB, Serruya M, Gepty A, Intenzo C, Lewis T, Amen D, et al. Clinical comparison of 99mTc exametazime and 123I Ioflupane SPECT in patients with chronic mild traumatic brain injury. PLoS One. 2014;9(1):1–9. doi: 10.1371/journal.pone.0087009

39. Khalili H, Rakhsha A, Ghaedian T, Niakan A MN. Application of brain perfusion SPECT in the evaluation of response to zolpidem therapy in consciousness disorder due to traumatic brain injury. 2020;(4): 315–320. doi: 10.4103/ijnm.IJNM_97_20

40. Nakagawara J, Kamiyama K, Takahashi M, Nakamura H. Cortical neuron loss in post-traumatic higher brain dysfunction using (123)I-iomazenil SPECT. Acta Neurochir Suppl. 2013;118:245–250. doi:10.1007/978-3-7091-1434-6_46

41. Womack KB, Dubiel R, Callender L, Dunklin C, Dahdah M, Harris TS, et al. (123)I-Iofluopane single-photon emission computed tomography as an imaging biomarker of pre-synaptic dopaminergic system after moderate-to-severe traumatic brain injury. J Neurotrauma. 2020;37(19):2113–2119. doi: 10.1089/neu.2019.6892

42. Raji CA, Henderson TA. PET and single-photon emission computed tomography in brain concussion. Neuroimaging Clin N Am. 2018;28(1):67–82. doi: 10.1016/j.nic.2017.09.003

43. Taipale J, Hautamäki L. Clinical practice guidelines in courts’ representation of medical evidence and testimony. Soc Sci Med. 2021;275. doi:10.1016/j.socscimed.2021.113805

44. Wortzel HS, Filley CM, Anderson CA, Oster T, Arciniegas DB. Forensic applications of cerebral single photon emission computed tomography in mild traumatic brain injury. J Am Acad Psychiatry Law. 2008;36(3):310–22.

45. Canadian Association of Nuclear Medicine (CANM). Guidelines for brain perfusion single photon emission computed tomography (SPECT); 2020. Available from: https://www.canm-acmn.ca/guidelines

46. Adinoff B, Devous M. Scientifically unfounded claims in diagnosing and treating patients. Am J Psychiatry. 2010;167(5):598. doi:10.1176/appi.ajp.2010.10020157

47. Amen D. Brain SPECT imaging in clinical practice. Am J Psychiatry. 2010;167(9):1125. doi:10.1176/appi.ajp.2010.10060814

48. Amen DG, Easton M. A New Way Forward: How brain SPECT imaging can improve outcomes and transform mental health care into brain health care. Front Psychiatry. 2021;12:1–17. doi:10.3389/fpsyt.2021.715315

49. Fisher M, Samuel F, Ping J, Ronald CK. Cerebral microbleeds in the elderly: A pathological analysis. Stroke. 2010;41(12):2782–2785. doi:10.1161/STROKEAHA.110.593657

50. Haller S, Vernooij MW, Kuijer JPA, Larsson EM, Jäger HR, Barkhof F. Cerebral microbleeds: Imaging and clinical significance. Radiology. 2018;287(1):11–28. doi: 10.1148/radiol.2018170803

